# Adjunctive Psychobiotic *Lactiplantibacillus plantarum* PS128 Therapy and Escitalopram in Major Depressive Disorder: A 12-Week Randomized, Double-Blind, Placebo-Controlled Trial

**DOI:** 10.64898/2026.08.25.26361081

**Authors:** Yunxin Ji, Jiale Zhang, Jieqiong Hu, Jiaxin Mao, Lanlan Wang, Kuilai Wang, Qingyu Zhang, Zhongze Lou, Yuwei Mi

**Affiliations:** The First Affiliated Hospital of Ningbo University, Zhejiang, China; The Fourth People’s Hospital of Haining, Jiaxing, China

**Author notes:** These authors contributed equally to this work (Yunxin Ji, Jiale Zhang). Corresponding author: Department of Psychosomatic Medicine, The First Affiliated Hospital of Ningbo University, Zhejiang, China. No. 59, Liutung Street, Haishu District, Ningbo City 315010, Zhejing, China. E-mail address: YJ; YM.

**Keywords:** Major depression disorder (MDD), serotonin selective reuptake inhibitor (SSRI), Lactobacillus plantarum PS128 (PS128), microbiota, HAMD

## Abstract

Major depressive disorder (MDD) is strongly associated with dysregulation of the hypothalamic-pituitary-adrenal (HPA) axis, systemic inflammation, and gut microbiota dysbiosis. Although selective serotonin reuptake inhibitors such as escitalopram are standard treatments, their efficacy is often constrained by partial response and gastrointestinal adverse effects. In this 12-week, randomized, double-blind, placebo-controlled trial, we evaluated the clinical efficacy and microecological mechanisms of adjunctive *Lactiplantibacillus plantarum* PS128 (PS128; 6 × 10^10^ CFU/day) in MDD patients on stable escitalopram therapy. Adjunctive PS128 significantly enhanced clinical response compared to placebo, yielding substantial reductions in HAMD-17 and MADRS, alongside a higher remission rate. 16S rRNA sequencing and PICRUSt2 profiling revealed that PS128 enriched key short-chain fatty acid producers (*Faecalibacterium*, *Coprococcus*), counteracting the *Klebsiella* expansion seen in placebo. Functionally, PS128 up-regulated neuroprotective cofactor, B vitamins, biosynthesis and down-regulated the neurotoxic kynurenine pathway. Network analysis demonstrated that PS128 maintained a resilient, integrated microbial co-occurrence topology, whereas the placebo network showed structural segregation. This stabilized ecosystem attenuated peripheral inflammatory signaling and normalized salivary cortisol levels. Overall, adjunctive PS128 augments escitalopram efficacy by enhancing gut network stability, supporting cellular energetics, and modulating neuroendocrine activity, offering a promising multimodal strategy for MDD.

**Trial Registration:** Chinese Clinical Trial Registry (ChiCTR2500101411).

**Highlights:**

- 12-week RCT assessed adjunctive PS128 in SSRI-treated patients with MDD.
- Adjunctive PS128 significantly improved depressive symptoms over SSRI alone.
- PS128 supplementation yielded a markedly higher clinical remission rate.
- PS128 stabilized gut microbiota topology and enriched key SCFA producers.
- PS128 upregulated bacterial B-vitamin biosynthesis and metabolic pathways.

## 1. Introduction

Major depressive disorder (MDD) represents a monumental global health challenge, affecting approximately 4.4% of the world’s population and ranking as a leading cause of disability worldwide (Santomauro et al., 2026; World Health Organization, 2017). In China, the epidemiological impact is also pronounced that the lifetime prevalence of MDD has reached 3.4–6.8%, impacting upwards of 54 million individuals and generating profound socioeconomic and healthcare burdens (Huang et al., 2019; Tian et al., 2026). Although standard pharmacological treatments—predominantly SSRIs such as escitalopram—remain the gold-standard therapeutic mainstays, their clinical utility is severely bottlenecked. Approximately 30–50% of patients fail to achieve adequate clinical response or full symptomatic remission under conventional first-line antidepressant monotherapy (Rush et al., 2006). Furthermore, the clinical administration of escitalopram is frequently complicated by treatment-associated somatic complaints, particularly gastrointestinal (GI) side effects including nausea, bloating, abdominal pain, and altered bowel habits, which drastically compromise patient adherence and overall treatment satisfaction (Oliva et al., 2021). This pervasive clinical ceiling of standard monotherapy, combined with secondary somatic distress, highlights an urgent mandate to explore alternative pathophysiological mechanisms and engineer targeted, well-tolerated adjunctive therapeutic strategies (Kennedy et al., 2009; Rush et al., 2006).

In recent years, the microbiota-gut-brain axis (MGBA) has emerged as a cornerstone bidirectional communication network that dynamically coordinates mood, cognition, and neuroendocrine homeostasis. This complex axis integrates neural (e.g., vagal signaling), neuroendocrine (the hypothalamic-pituitary-adrenal [HPA] axis), systemic immune (circulating cytokines), and microbial metabolic channels to translate peripheral intestinal ecology into central nervous system (CNS) function (Cryan et al., 2019; Morais et al., 2021).

Accumulating clinical evidence demonstrates that individuals with MDD exhibit characteristic gut microbial dysbiosis, typically marked by reduced α-diversity (Lin et al., 2025), depletion of anti-inflammatory and short-chain fatty acids (SCFAs)-producing beneficial taxa (such as *Coprococcus*, *Blautia*, and *Faecalibacterium*), and enrichment of opportunistic pathogens (such as *Bilophila*, *Collinsella*, and *Dorea*)(Jiang et al., 2015; Radjabzadeh et al., 2022; Valles-Colomer et al., 2019). These microbial perturbations may further disrupt intestinal barrier integrity—often referred to as “leaky gut”—facilitating the translocation of bacterial components and metabolites into systemic circulation. This translocation triggers subclinical systemic inflammation, elevates pro-inflammatory cytokines (such as interleukin-6 [IL-6] and tumor necrosis factor-alpha [TNF-α]), and drives neuroinflammation and HPA axis hyperactivity (Hu et al., 2023; Liu et al., 2023, 2024). Additionally, the gut microbiota actively modulates host neurotransmitter synthesis by producing or regulating neuroactive compounds, including serotonin, dopamine, and gamma-aminobutyric acid, further establishing its critical role in depression pathogenesis (Sivamaruthi et al., 2026; Strandwitz, 2018).

Consequently, targeting the MGBA via probiotics—specifically “psychobiotics” defined as live microorganisms that confer mental health benefits when ingested in adequate amounts—has attracted significant research interest (Dinan et al., 2013). Among these, *Lactiplantibacillus plantarum* PS128 (formerly *Lactobacillus plantarum* PS128; PS128), a unique psychobiotic strain isolated from traditional Taiwanese fermented mustard greens (“Fu-tsai”)(Chao et al., 2009), has demonstrated exceptional psychotropic potential. In preclinical murine models, oral administration of PS128 successfully modulated hyperactive HPA axis signaling (reducing serum corticosterone), neutralized peripheral inflammatory cascades (decreasing IL-6), and balanced monoamine neurotransmitters (serotonin and dopamine) within the striatum and prefrontal cortex, thereby reversing chronic stress-induced anxious and depressive behaviors (W.-H. Liu et al., 2016; Y.-W. Liu et al., 2016). Clinically, PS128 supplementation has shown efficacy in alleviating stress, anxiety, and depressive symptoms and improving sleep quality in highly stressed professionals (Wu et al., 2021) and individuals with self-reported insomnia (Ho et al., 2021). It has also been shown to improve social communication and attention in children with neurodevelopmental disorders, such as autism spectrum disorder (ASD)(Liu et al., 2019).

Despite these promising findings, the clinical efficacy of PS128 in patients with clinically diagnosed MDD remains controversial. In a preliminary 8-week open-label trial involving 11 patients with MDD, the authors observed that adjunctive PS128 significantly decreased depressive scores (Chen et al., 2021). However, a subsequent rigorous 8-week randomized, double-blind, placebo-controlled trial failed to detect statistically significant superiorities in depression severity (measured by HAMD-17 and DSSS), systemic inflammatory profiles, gut permeability biomarkers, or macro-level gut microbiota architecture between the PS128 adjunct arm and the placebo arm, both group showed improvement in depression symptoms (Lin et al., 2024). This therapeutic divergence reveals a profound knowledge gap in psychobiotic science: the temporal kinetics of microecological rewiring. Emerging neuropsychopharmacological paradigms suggest that a severely dysbiosis-resilient gut microbiome in moderate-to-severe MDD possesses a high degree of microecological inertia; consequently, a standard 8-week intervention window may fundamentally insufficient to overcome this biological resistance, establish stable metabolic cross-feeding, and achieve the systemic neuroendocrine tuning required to translate peripheral changes into hard clinical efficacy (Alli et al., 2022; Cryan et al., 2019; Valles-Colomer et al., 2019). Overcoming this “therapeutic lag” may strictly require a more sustained longitudinal intervention threshold.

To address these challenges, we designed and conducted a 12-week, randomized, double-blind, placebo-controlled clinical trial. We recruited patients with MDD who were undergoing stable escitalopram treatment and randomized them to receive either adjunctive PS128 or a matching placebo. By extending the intervention period to 12 weeks, we aimed to overcome the temporal threshold limitations of previous 8-week designs. Our primary objective was to rigorously evaluate the superiority of adjunctive PS128 over standard escitalopram monotherapy (placebo group) in accelerating the reduction of core depressive symptoms and mitigating concomitant somatic GI distress. Our secondary objectives were to map the longitudinal neuroendocrine dynamics of the HPA axis (via salivary cortisol) and to comprehensively chart the structural and functional metagenomic modifications within the gut microbiota, thereby providing an empirical multi-omic framework for pharmacological-microbiological synergy in psychiatric management.

## 2. Materials and Methods

### 2.1. Ethical Approvals and Trial Registration

This study was approved by the Medical Ethics Committee of the First Affiliated Hospital of Ningbo University (Ningbo, China; 2023-R003-01). The clinical trial protocol, designed and conducted in strict accordance with the Declaration of Helsinki. All participants provided written informed consent prior to any study-related procedures. The study was retrospectively registered at Chinese Clinical Trial Registry (ChiCTR2500101411).

### 2.2. Participant Recruitment and Selection Criteria

Participants were consecutively recruited from the outpatient psychiatric clinic at the First Affiliated Hospital of Ningbo University between July 2023 and December 2024. Eligible participants met the following inclusion criteria: (1) aged between 18 and 65 years; (2) met the Diagnostic and Statistical Manual of Mental Disorders, Fifth Edition (DSM-5) or International Classification of Diseases, Tenth Revision (ICD-10) criteria for a major depressive episode; (3) had a 17-item Hamilton Depression Rating Scale (HAMD-17) score ≥18 (and/or ≥14); and (4) were first-episode or medication-naïve patients. Furthermore, participants must have been on a stable, fixed dose of escitalopram (10 mg/day) for at least 3 months prior to randomization, with this dosage maintained throughout the 12-week trial.

Exclusion criteria included: (1) comorbid psychiatric diagnoses (e.g., schizophrenia, bipolar disorder, or organic mental disorders); (2) significant systemic medical conditions, such as inflammatory bowel disease, autoimmune disorders, or diabetes; (3) active suicidal or homicidal ideation (defined as a score ≥3 on the HAMD-17 suicide item); (4) treatment-resistant depression or a history of receiving electroconvulsive therapy; (5) history of serious adverse events or hypersensitivity to escitalopram; (6) consumption of prescribed antibiotics, probiotics, prebiotics, or immunosuppressants within 4 weeks (1 month) prior to enrollment; and (7) pregnancy, lactation, or substance abuse.

### 2.3. Sample Size Calculation

The sample size calculation was based on the pilot open-label trial data from Chen et al. (Chen et al., 2021). In that study, the mean HAMD-17 score of patients with MDD was 20.1 (SD = 5.6) before PS128 treatment and decreased to 12.0 (SD = 6.1) post-treatment. The mean score of the control group was assumed to decrease to 18.0 (SD = 5.8). Based on an independent-samples t-test, the estimated effect size (Cohen’s d) was 0.8. With a significance level (α) of 0.05 and a statistical power (1-β) of 0.80, the required sample size was calculated using the formula:

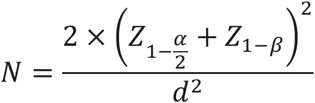

This calculation yielded a total required sample size of 50 participants (25 participants per group). Factoring in an anticipated 10% dropout rate, a final target sample size of 56 participants (28 participants per group) was determined for enrollment.

### 2.4. Randomized Double-Blind Place-Controlled Trial Design

This was a 12-week, single-center, randomized, double-blind, placebo-controlled parallel-group trial. Eligible participants were randomly assigned in a 1:1 ratio to receive either adjunctive psychobiotic PS128 or a matching placebo. Randomization was conducted using block randomization with computer-generated random numbers. The allocation sequence was managed by an independent pharmacist and sealed in sequentially numbered, opaque, identical bottles to maintain strict blinding. All participants, clinical investigators, and outcome assessors remained blinded to the treatment assignment until the final database lock and unblinding.

### 2.5. Pharmacological and Psychobiotic Interventions

All participants received standard escitalopram oxalate tablets (Lepeng, Zhejiang Huahai Pharmaceutical Co., Ltd., China; 10 mg/tablet) as the background antidepressant. The starting dose of escitalopram was 5 mg/day (taken orally as half a tablet in the morning) for the first 7 days, which was then increased to 10 mg/day (1 tablet in the morning) on Day 8. Depending on clinical response and tolerability, the dose could be adjusted up to a maximum of 20 mg/day (2 tablets in the morning) after 14 days. All medications were consumed 30 minutes after meals. Alongside the escitalopram background, participants were asked to take 2 study capsules, probiotic or placebo, 30 minutes after dinner. The study capsules were provided by Asian Probiotics and Prebiotics Corporation (Shanghai, China). The probiotic capsule contains *Lactiplantibacillus plantarum* PS128 (≥3×10^10^ CFU/capsule) providing live PS128 with daily dose of 6×10^10^ CFU. The placebo capsule was visually, physically, and organoleptically identical to probiotic one. The placebo capsules contain microcrystalline cellulose (300 mg/capsule). Both groups underwent intervention for 12 weeks. Concomitant use of other antidepressants, antipsychotics, mood stabilizers, antibiotics, or other probiotic/prebiotic supplements was strictly prohibited during the trial. If insomnia occurred, non-benzodiazepine hypnotics were permitted for a maximum of 2 weeks based on clinical necessity.

### 2.6. Clinical Outcome Assessments

Clinical visits and psychiatric evaluations were scheduled at baseline (V1), Week 4 (V2), Week 8 (V3), and Week 12 (V4). Core clinician-rated scales included the HAMD-17 (to evaluate objective depressive severity)(Hamilton, 1960), the Montgomery-Åsberg Depression Rating Scale (MADRS) (to sensitively capture core mood swings)(Montgomery and Åsberg, 1979), and the Clinical Global Impression-Severity (CGI-S) and Clinical Global Impression-Improvement (CGI-I) scales (to rate overall disease severity and relative improvement)(William Guy, 1976). Patient-reported self-rating instruments included the Patient Health Questionnaire-9 (PHQ-9) (for subjective depressive severity)(Kroenke et al., 2001), the Gastrointestinal Symptom Rating Scale (GSRS) (to evaluate bowel function and abdominal distress)(Svedlund et al., 1988), and the Patient Global Impression-Improvement (PGI-I) scale (for subjective efficacy perception)(Yalcin and Bump, 2003). Clinician-rated scales were completed via structured, one-on-one psychiatric interviews conducted by certified clinical evaluators. Patient self-reports were gathered using paper or digital questionnaires.

### 2.7. Fecal DNA Extraction and 16S rRNA Amplicon Sequencing

Fresh fecal specimens were collected at baseline (V1) and Week 12 (V4). Fecal samples were deposited into sterile containers, temporarily frozen at −20 °C for a maximum of 7 days, and subsequently transported on dry ice to the laboratory for long-term storage at −80 °C until DNA extraction. Fecal DNA was extracted from 0.2 g of fecal sample using the FastPure Stool DNA Isolation Kit (Vazyme Biotech, Nanjing, China) following the manufacturer’s protocol. The concentration and purity of the extracted genomic DNA were verified using a NanoDrop 2000 spectrophotometer (Thermo Fisher Scientific, Waltham, MA, USA), and DNA integrity was checked via 1% agarose gel electrophoresis. The hypervariable V3-V4 region of the bacterial 16S rRNA gene was amplified using the universal primers 338F (5’-ACTCCTACGGGAGGCAGCAG-3’) and 806R (5’-GGACTACHVGGGTWTCTAAT-3’). PCR amplifications were performed in triplicate in a 20 μL reaction system containing: 4 μL of 5x FastPfu Buffer, 2 μL of dNTPs (2.5 mM), 0.8 μL of each primer (5 μM), 0.8 μL of reverse primer (5 μM), 0.4 μL of FastPfu DNA Polymerase, 0.2 μL of BSA, 10 ng of template DNA, and sterile ddH_2_O to reach the final volume of 20 μL. The PCR was conducted on ABI GeneAmp 9700 with the amplification program: initial denaturation at 95 °C for 3 min; 30 cycles of denaturation at 95 °C for 30 s, annealing at 53 °C for 30 s, and extension at 72 °C for 45 s; followed by a final extension step at 72 °C for 10 min. The triplicate PCR products were pooled, evaluated on a 2% agarose gel, and purified using a AMPure® PB beads (Pacifc Biosciences, CA, USA). The purified PCR products were quantified using a QuantiFluor™ –ST (Promega) and mixed in equimolar ratios. High-throughput sequencing libraries were prepared using the NEXTFLEX® Rapid DNA-Seq Kit. Paired-end sequencing (2 × 300 bp) was performed on the Illumina NextSeq 2000 platform at Majorbio Bio-Pharm Technology Co. Ltd. (Shanghai, China).

### 2.7. Salivary Cortisol Processing and Quantification

Saliva samples were collected from participants at 1:00 PM during each visit (V1 to V4). Participants were instructed to sit quietly for 10 minutes prior to sampling and to refrain from brushing their teeth or drinking water during this pre-sampling period. Saliva was collected using standard saliva collection tubes. The collected specimens were centrifuged at 3000 r/min for 10 minutes at 4 °C to separate the cellular debris. The supernatant was immediately aliquoted and stored in cryovials at −20 °C until analysis. Salivary cortisol levels were quantitatively determined using a high-sensitivity enzyme-linked immunosorbent assay (ELISA) kit (Human Salivary Cortisol ELISA Kit, KT111793-A; Jiangsu Kete Biotechnology Co., Ltd., China) following the manufacturer’s instructions. Absorbance (OD value) was measured, and a linear regression standard curve was plotted in Excel to calculate the actual cortisol concentration (nmol/L) for each sample.

### 2.8. Bioinformatics Processing and Statistical Analysis

Raw sequencing reads were quality-controlled using fastp software (v0.19.6) to filter low-quality bases, adapter sequences, and reads shorter than 50 bp. Overlapping reads were assembled using FLASH (v1.2.7) with a minimum overlap of 10 bp and a maximum mismatch of 20%. Assembled sequences were denoised and clustered into Amplicon Sequence Variants (ASVs) using the DADA2 plugin within QIIME2 (v2021.4)(Bolyen et al., 2019). Chloroplast– and mitochondria-derived ASVs were removed. To normalize sequencing depth, all samples were rarefied to a uniform depth of 19,234 sequences per sample, achieving an average Good’s coverage of 99.99%. Taxonomic classification was performed using the QIIME2 Naive Bayes classifier trained against the Silva 16S rRNA database (v138.1). Alpha diversity metrics (including Chao1 richness, Shannon diversity, and Faith_PD) were calculated using phyloseq (v1.48.0) and compared longitudinally and between groups using Wilcoxon signed-rank tests. Beta diversity was assessed based on Bray-Curtis distance matrices, visualized using Principal Coordinate Analysis (PCoA), and analyzed using Permutational Multivariate Analysis of Variance (PERMANOVA) with 9,999 permutations. MaAsLin2 (Mallick et al., 2021) and Linear Discriminant Analysis Effect Size (LEfSe)(Segata et al., 2011) was employed to detect biomarkers at the genus level with an LDA score threshold > 2 and P<0.05 in R (Maaslin2 v1.18.0). Gut microbiota functional pathways were predicted using PICRUSt2 (Douglas et al., 2020) against the KEGG and MetaCyc databases on Galaxy Europe (https://usegalaxy.eu/)(v2.3.0-b). Demographic and clinical measured data were statistically analyzed using R (version 4.4.1) in RStudio (2025.09.2). Generalized estimating equations (GEE; geepack v1.3.13) and LMM (lmerTest v2.0-1) were applied to analyze longitudinal trajectories, adjusting for baseline scores, age, and gender. Group-by-time interactions were evaluated. Group differences in demographics and baseline clinical variables were compared using Student’s t-test or chi-square test. Spearman correlation was used to determine the relationships between taxonomic abundance and clinical scores. P-values were adjusted using the Benjamini-Hongberg FDR method for multiple comparisons. To elucidate the functional co-occurrence interactions between key microbial taxa and predicted metabolic pathways, microbial co-occurrence network analysis was performed for both the Probiotic and Placebo groups. The non-parametric Spearman’s rank correlation coefficients (rho) were calculated based on the relative abundances of differentiated bacterial genera and PICRUSt2-predicted KEGG pathways.

Correlations with |rho| >0.4 and FDR < 0.05 were retained to construct the network edges. The filtered correlation matrices were subsequently imported into Gephi (version 0.11.2) for topological visualization and analysis. Node size was proportional to its betweenness centrality, calculated via the Network Diameter algorithm with normalized values [0, 1], to highlight the critical functional hubs. Network modularity was computed using the Louvain algorithm with a resolution setting of 1.0 to partition the network into distinct functional clusters (Modularity Classes). The overall graph spatial arrangement was optimized using the ForceAtlas 2 layout algorithm, followed by minor manual adjustments to prevent label occlusion and ensure visual comparability between the two arms.

## 3. Results

### 3.1. Study Cohort, Participant Flow, and Baseline Demographics

A total of 62 patients with major depressive disorder (MDD) receiving stable escitalopram therapy were enrolled and randomized into either the adjunctive PS128 group (n = 32) or the placebo group (n = 30). Baseline demographic and clinical characteristics were evaluated in the full randomized population (n = 62) and were comparable between the two arms (all P > 0.05; **Table 1**). Over the 12-week intervention, a total of 9 participants discontinued the study. In the PS128 group, 3 participants dropped out prior to Week 4 due to refusal to continue (n = 2) and loss to follow-up (n = 1), leaving 29 participants who completed the 12-week protocol. In the placebo group, 6 participants discontinued intervention due to capsule dissatisfaction (n = 2; Week 4), refusal to continue (n = 2; Week 4), and loss to follow-up (n = 2; Week 8 and Week 12), resulting in 24 participants completing the trial (**Figure 1**). Consequently, the per-protocol (PP) population comprised 53 participants (PS128: n = 29; placebo: n = 24) with complete clinical evaluations across all study visits and paired fecal samples at baseline and Week 12.

**Figure 1.**
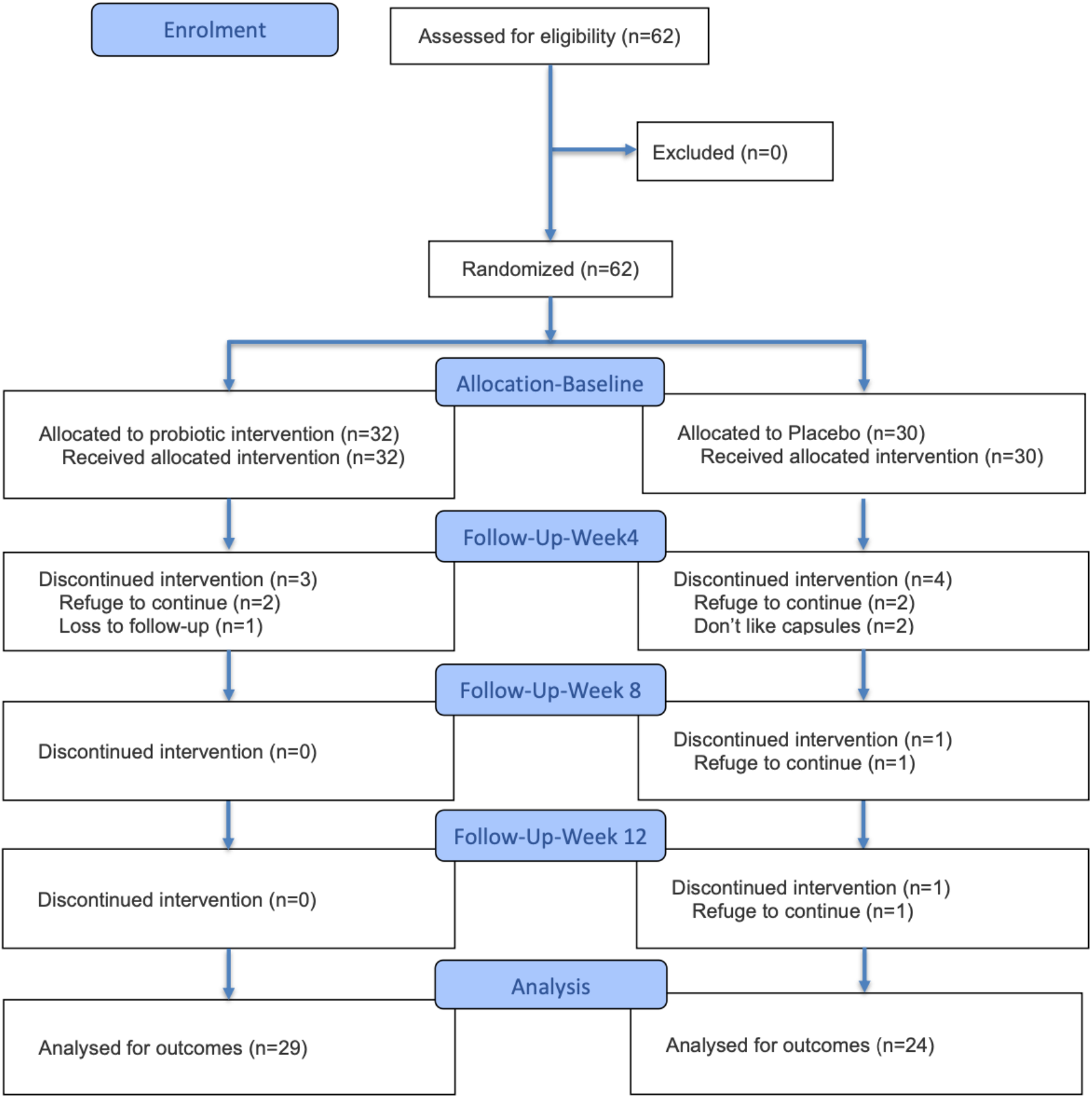
CONSORT flow diagram of participant progress through the trial. Schematic representation of participant screening, randomization, 12-week intervention allocation, follow-up, and analytical cohorts in a randomized, double-blind, placebo-controlled trial evaluating adjunctive *Lactiplantibacillus plantarum* PS128 in patients with major depressive disorder (MDD) receiving stable escitalopram therapy. A total of 62 eligible participants were randomized in a 1:1 ratio to receive either adjunctive PS128 (n=32) or matching placebo (n=30), forming the baseline randomized population (n=62). Over the 12-week study period, 3 participants in the PS128 group discontinued prior to Week 4 (refusal to continue, n=2; loss to follow-up, n=1), and 6 participants in the placebo group discontinued intervention due to capsule dissatisfaction (n = 2; Week 4), refusal to continue (n = 2; Week 4), and loss to follow-up (n = 2; Week 8 and n =2; Week 12). A total of 53 participants completed all study visits with paired biological samples (PS128: n=29; placebo: n=24) and were included in the final per-protocol (PP) analysis for clinical and multi-omic outcomes.

**Table 1.**
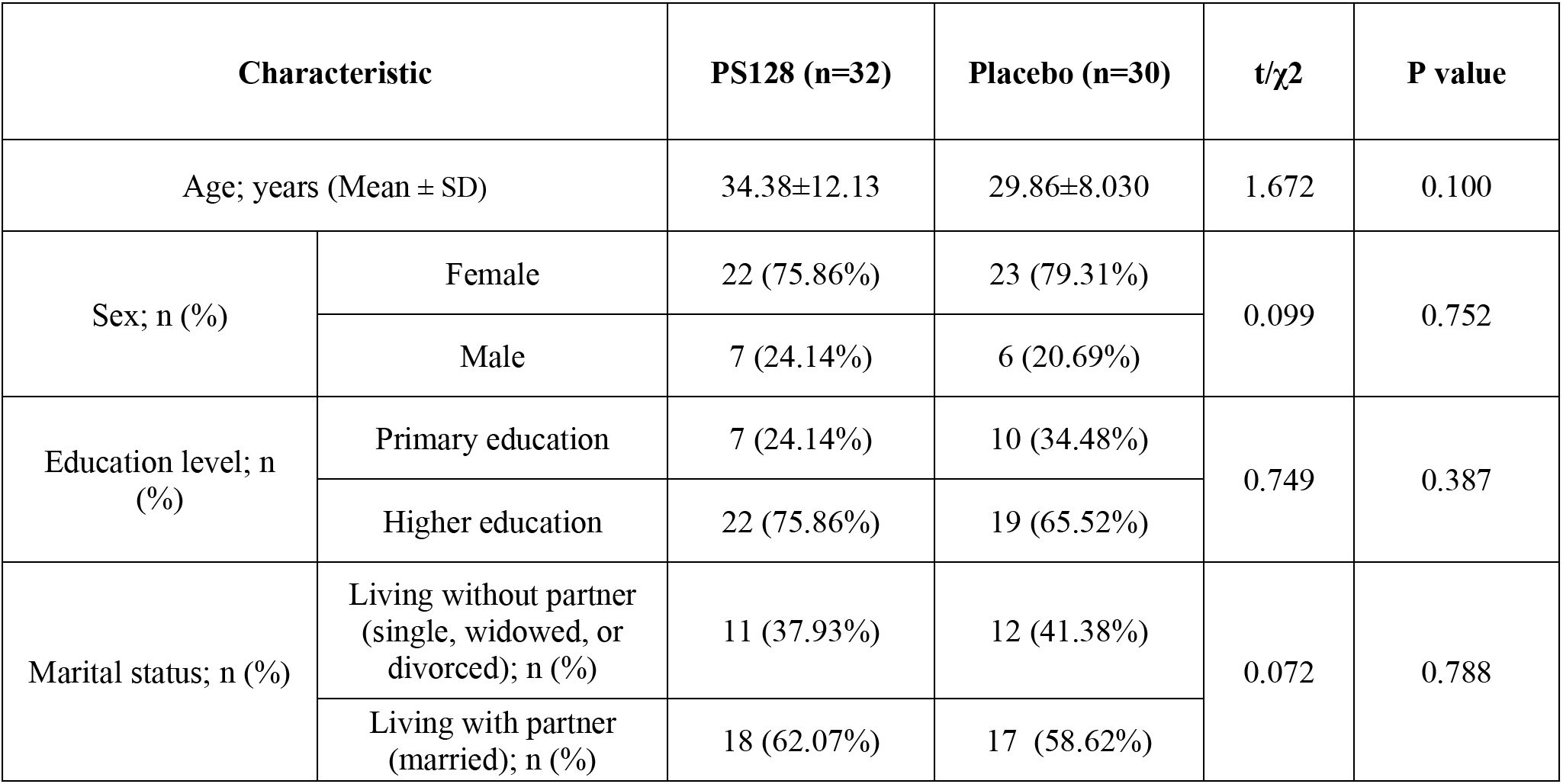
Baseline demographic and clinical characteristics of the study participants. Comparison of demographic variables (age, sex, education level, and marital status) and baseline clinical severity scores (HAMD-17, MADRS, PHQ-9, and GSRS) between the adjunctive PS128 group (n = 32) and the placebo group (n = 30) randomized at V1. Continuous variables are expressed as mean ± SD and compared using independent Student’s t-tests or Mann-Whitney U tests. Categorical variables are presented as frequencies (percentages) and evaluated via Pearson’s chi-square test or Fisher’s exact test. P > 0.05 indicates no statistically significant differences between groups, establishing baseline homogeneity.

| Characteristic | | PS128 (n=32) | Placebo (n=30) | t/$\chi^2$ | P value |
| --- | --- | --- | --- | --- | --- |
| Age; years (Mean ± SD) | | 34.38±12.13 | 29.86±8.030 | 1.672 | 0.100 |
| Sex; n (%) | Female | 22 (75.86%) | 23 (79.31%) | 0.099 | 0.752 |
|  | Male | 7 (24.14%) | 6 (20.69%) |  |  |
| Education level; n (%) | Primary education | 7 (24.14%) | 10 (34.48%) | 0.749 | 0.387 |
|  | Higher education | 22 (75.86%) | 19 (65.52%) |  |  |
| Marital status; n (%) | Living without partner (single, widowed, or divorced); n (%) | 11 (37.93%) | 12 (41.38%) | 0.072 | 0.788 |
|  | Living with partner (married); n (%) | 18 (62.07%) | 17 (58.62%) |  |  |

### 3.2. Efficacy of Adjunctive PS128 on Primary Depressive Symptoms

Over the 12-week intervention, both the adjunctive probiotic group and the placebo group (representing escitalopram monotherapy) showed significant within-group reductions in all primary depressive scales, including the clinician-rated HAMD-17, MADRS, and the patient-reported PHQ-9 (all *P* < 0.05 versus baseline V1; **Figure 2A**, **Table S1**). However, LMM revealed highly significant group-by-time interactions. The adjunctive probiotic PS128 group exhibited significantly faster and more pronounced symptom reduction than the placebo group. Post-hoc pairwise comparisons using estimated marginal means (EMMs) with Bonferroni correction further pinpointed the temporal onset of this therapeutic efficacy (**Table S2**). While no significant baseline difference in HAMD-17 scores was observed between the two groups at V1 (*P* > 0.05), the probiotic group demonstrated a significantly lower clinical score compared to the placebo group starting from V3 (*P* = 0.024), an advantage that became even more pronounced by the V4 endpoint (*P* < 0.001). At the Week 12 (V4) endpoint; the probiotic group dropped to mild-to-subclinical states, whereas the placebo group remained at moderate severity (*P* < 0.0001), demonstrating a remarkably strong effect size (Cohen’s d = −1.052, 95% CI: –1.643 to –0.461). MADRS scores displayed a highly similar pattern of superiority for the PS128 adjunct at Week 12 (Cohen’s d=−1.067, 95% CI: –1.659 to –0.475). Subjective depressive burdens evaluated by PHQ-9 also showed significantly lower scores in the probiotic group at Week 12 (P < 0.05, Cohen’s d=−0.537, 95% CI: –1.101 to 0.211). The detailed longitudinal parameters of all clinical measurements are consolidated in **Figure 2** and **Table S1**.

**Figure 2.**
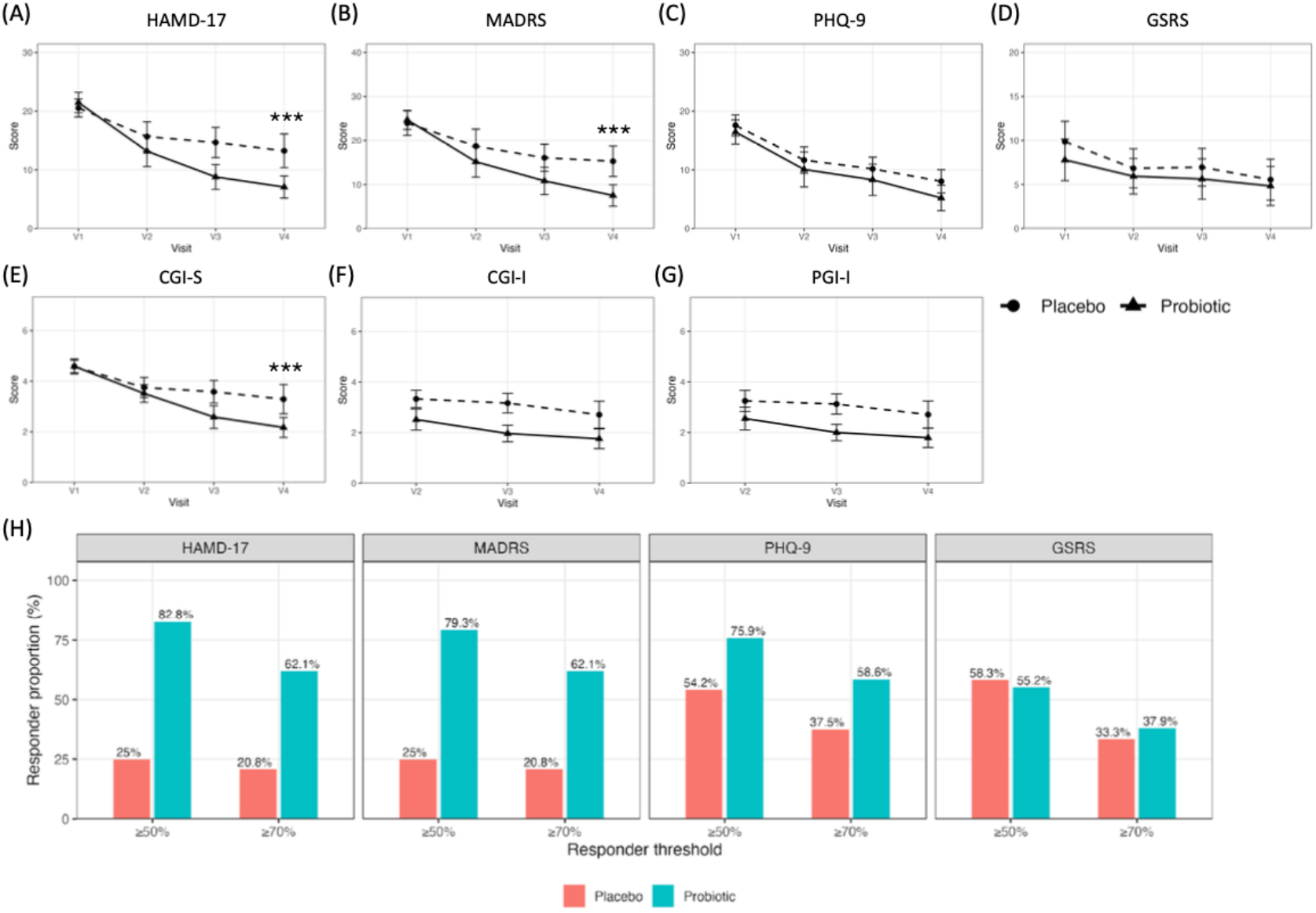
Efficacy of adjunctive PS128 therapy on primary depressive, global clinical, and gastrointestinal outcomes. Longitudinal trajectory and treatment outcomes over the 12-week intervention. (A) 17-item Hamilton Depression Rating Scale (HAMD-17) scores across study visits. (B) Montgomery-Åsberg Depression Rating Scale (MADRS) scores. (C) Patient Health Questionnaire (PHQ-9) scores. (D) Gastrointestinal Symptom Rating Scale (GSRS) scores. (E–G) Clinical Global Impression-Severity/Improvement (CGI-S/I) and Patient Global Impression-Improvement (PGI-I) score distributions. (H) Clinical response (defined as a ≥ 50% score reduction from baseline) and clinical remission (defined as a ≥ 70% score reduction) rates at Week 12. Data are presented as mean ± SEM or percentages. Statistical significance for longitudinal trajectories was evaluated using Linear Mixed-Effects Models (LMM) followed by post-hoc pairwise comparisons with Bonferroni correction. *P < 0.05, ** P < 0.01, and *** P < 0.001, indicate statistically significant differences between the PS128 and placebo groups at specific time points.

### 3.3. Secondary Outcomes: Gastrointestinal Symptoms and Clinical Global Impressions

Subjective gastrointestinal distress was measured using the GSRS (**Figure 2D**). At baseline, the probiotic group exhibited non-significantly higher GSRS scores than the placebo group (*P* = 0.16), suggesting a slightly higher burden of baseline gastrointestinal comorbidities. Following the 12-week intervention, the PS128 group showed a significant within-group reduction in GSRS scores (*P* < 0.05), whereas the placebo group demonstrated no significant change (*P* > 0.05). However, the inter-group difference at Week 12 did not reach statistical significance (Cohen’s d = −0.125, *P* = 0.43), indicating that the therapeutic benefits of PS128 were predominantly centered on psychiatric symptoms rather than peripheral GI distress. For the Global Impressions (**Figure 2E-G**), CGI-S scores decreased in both cohorts, with the PS128 group achieving a significantly lower severity level at Week 12 than the placebo group (*P* < 0.0001, Cohen’s d = −0.935). Consistently, both clinician-rated (CGI-I) and patient-rated (PGI-I) improvement scores were significantly lower—indicating greater clinical improvement—in the PS128 group compared to the placebo group from Week 4 through Week 12 (*P* < 0.001, Cohen’s d = −0.833 at Week 12). Furthermore, Pearson correlation analysis revealed a highly significant positive correlation between CGI-I and PGI-I scores (*r* = 0.978, *P* < 0.0001), underscoring a strong robust agreement between clinician assessment and patient self-perception (**Table S2**).

### 3.4. Response and Remission Proportions

To evaluate the clinical importance of PS128 as adjunct to escitalopram in MDD, participant improvements were stratified according to clinical response (defined as a ≥50% reduction from baseline scores) and clinical remission (defined as a ≥70% reduction). As shown in **Figure 2H**, at Week 12, the clinical response rate on the primary HAMD-17 scale reached 82.8% in the PS128 group compared to only 25.0% in the placebo group. More importantly, a high clinical remission rate (≥70% HAMD-17 reduction) was achieved by 62.1% of patients in the PS128 group, which was significantly greater than the 20.8% observed in the placebo group. For the MADRS scale, the response and remission rates in the PS128 group were 79.3% and 62.1%, respectively, compared to 25.0% and 20.8% in the placebo group. On the self-reported PHQ-9, 75.9% and 58.6% of patients in the PS128 group reported response and remission, respectively, compared to 37.5% and 33.3% in the placebo group. These outcomes collectively demonstrate that the adjunctive administration of PS128 substantially increased the likelihood of achieving profound and clinically meaningful symptom remission compared to escitalopram as standard antidepressant monotherapy.

### 3.5. Gut Microbiota Alpha and Beta Diversity Trajectories

To evaluate the impact of PS128 on the gut microecology, 16S rRNA gene sequencing was conducted on fecal samples collected at baseline (V1) and Week 12 (V4). Standardized to an even depth of 19,234 reads, the α-diversity indices are comparable between two groups at V4. The placebo group showed no significant changes in any α-diversity indices over the 12-week trial. In contrast, the adjunctive PS128 group exhibited significant increases in species richness and evenness (**Figure 3A**). The Shannon diversity index significantly increased from V1 to V4 in the PS128 group (paired Wilcoxon signed-rank test, *P*_adj_=0.008, ES=0.871). This expansion of diversity was also observed in Chao1 species richness (from V1 to V4, *P*_adj_=0.008, ES=0.665) and Faith’s Phylogenetic Diversity index (Faith_PD: *P*_adj_=0.008, ES=0.671). Beta-diversity Principal Coordinate Analysis (PCoA) further confirmed the structural remodeling of the gut microbiota (**Figure 3B**). Under the Bray-Curtis distance matrix, PERMANOVA revealed a highly significant shift in microbial community structure between the groups at Week 12 (P=0.003). Jaccard distance metrics, which reflect species presence/absence, showed only a marginal shift (*P* = 0.075), suggesting that PS128 primarily modulated the relative abundances of existing commensal taxa rather than inducing a complete species turnover. Furthermore, PCoA spatial analysis revealed that the PS128 group showed significantly longer longitudinal migration distances (Euclidean distance in PCoA space) from V1 to V4 than the placebo group, illustrating robust individual-level microbial community shifts.

**Figure 3.**
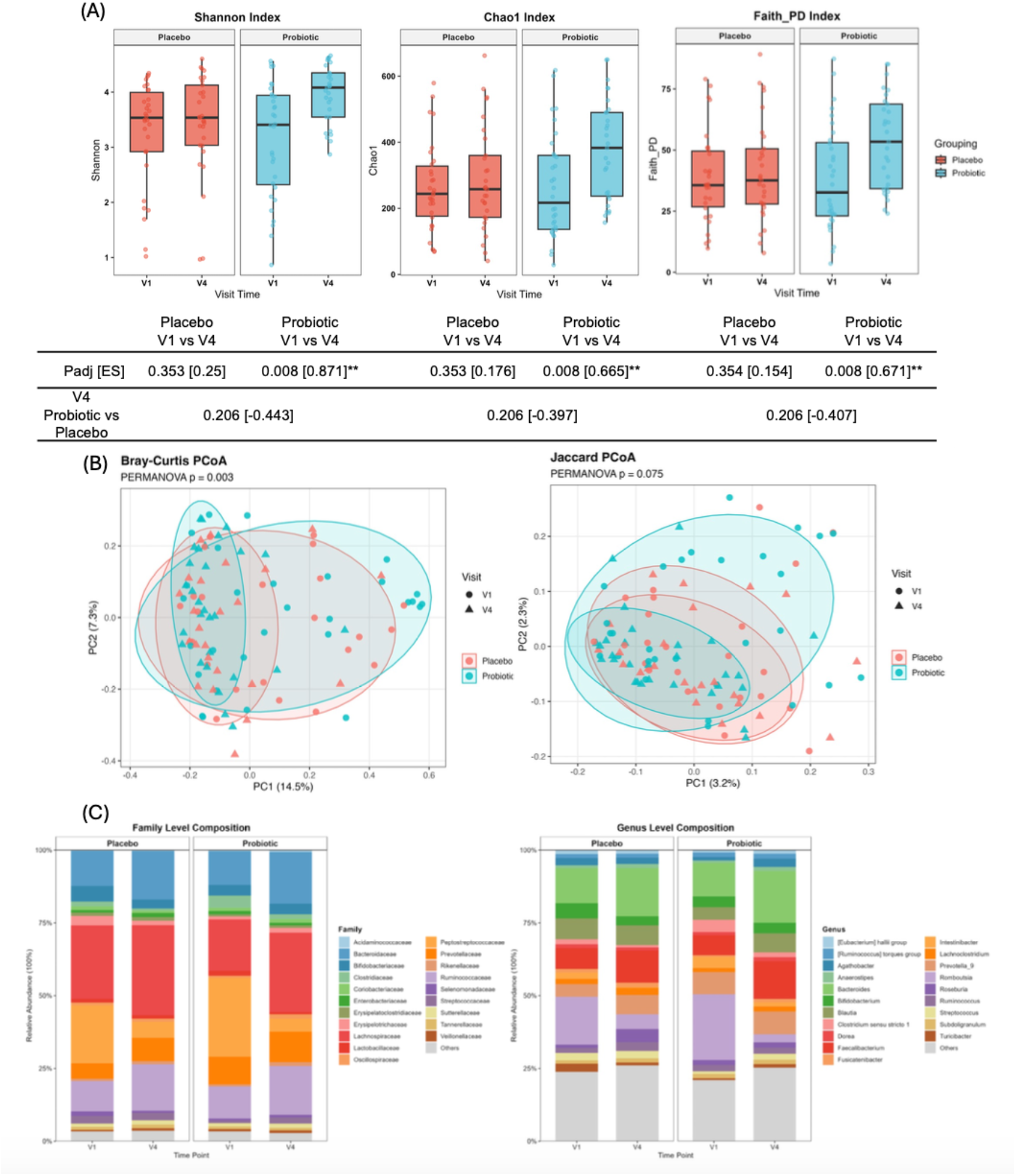
Alterations in gut microbiota alpha and beta diversity trajectories. (A) Within-individual α-diversity dynamics quantified by Shannon, Chao1, and Faith’s Phylogenetic Diversity (Faith_PD) indices from baseline (V1) to Week 12 (V4) for the PS128 and placebo groups. (B) β-Diversity Principal Coordinate Analysis (PCoA) based on the Bray-Curtis dissimilarity matrix, demonstrating structural community shifts between groups at Week 12. Longitudinal *α*-diversity changes were evaluated using paired Wilcoxon signed-rank tests with Benjamini-Hochberg false discovery rate correction (P_adj_). Between-group β-diversity divergence was tested using Permutational Multivariate Analysis of Variance (PERMANOVA; 9,999 permutations).

### 3.6. Microbial composition and taxonomic shifts

Taxonomic annotation at the family and genus levels revealed broadly similar baseline microbial profiles between the Placebo and Probiotic groups (**Figure 3C, D**). To detect specific biomarker shifts, differential abundance analysis was performed using both LEfSe and MaAsLin2 for cross-validation (**Figure 4**). At the 12-week endpoint (V4), LEfSe identified a significant enrichment of the class *Verrucomicrobiae*, the family *Lactobacillaceae*, and the genera *Lactiplantibacillus*, *Agrilactobacillus*, and *Akkermansia* in the Probiotic group, whereas the genus *Klebsiella* was significantly enriched in the Placebo group (**Figure 4A**).

**Figure 4.**
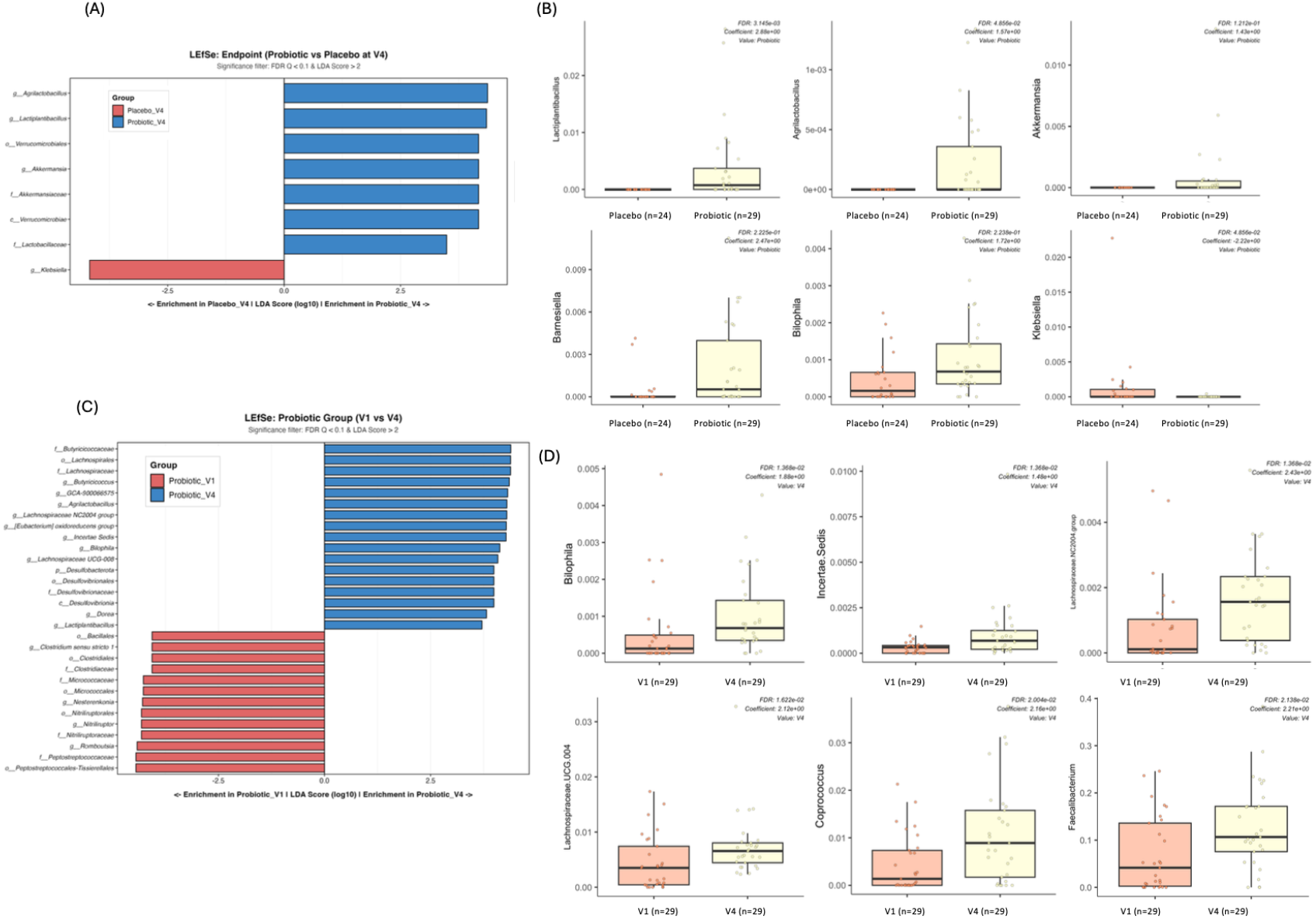
Taxonomic biomarker profiling and clinical correlation analysis. (A) Linear Discriminant Analysis Effect Size (LEfSe) and (B) Multivariate Association with Linear Models (MaAsLin2) identifying differential taxonomic features enriched in the PS128 vs. placebo groups at Week 12 (LDA score > 2.0, P < 0.05, FDR <0.05). (C) Relative abundance profiles of key short-chain fatty acid (SCFA)-producing and psychobiotic genera across treatment arms. (D) Heatmap of Spearman rank correlation coefficients between differential microbial abundance and clinical rating scores (HAMD-17, MADRS, PHQ-9, GSRS). Red indicates positive correlation; blue indicates negative correlation. * P <0.05, ** P <0.01.

Complementing these findings, MaAsLin2 analysis confirmed these probiotic-induced taxonomic shifts; it uniquely captured additional significant enrichments of *Barnesiella* and *Bilophila* in the probiotic-treated cohort relative to placebo at V4 (**Figure 4B**). Longitudinal intra-group comparisons within the Probiotic cohort (V1 vs. V4) were subsequently conducted to evaluate temporal shifts. LEfSe analysis revealed a post-intervention enrichment of several SCFA-producing taxa, including *Butyricicoccus* and *Dorea* (**Figure 4C**). Furthermore, MaAsLin2 identified significant increases in the abundance of *Bilophila*, *Coprococcus*, and *Faecalibacterium*, alongside unclassified bacterial taxa, following the 12-week adjunctive PS128 intervention (**Figure 4D**).

To systematically decode the functional landscape and microbial cross-talk driving the observed clinical outcomes, we combined predictive metagenomic profiling with higher-order topological network analyses (**Figures 5A–5D**). At the macro-functional level, MetaCyc pathway differentiation via PICRUSt2 revealed a profound divergence between the two groups that 30 functional pathways diverged significantly between the Placebo and Probiotic groups post-intervention (**Figure 5A**). The Probiotic group (PS128 adjunctive to escitalopram) exhibited a coordinated enrichment in critical cofactor and vitamin biosynthetic machineries, including thiamine diphosphate biosynthesis I (THISYN-PWY; Vitamin B1), 6-hydroxymethyl-dihydropterin diphosphate biosynthesis III (PWY-7539; tetrahydrofolate biosynthesis and salvage), and phosphopantothenate biosynthesis I (PANTO-PWY; pantothenate synthesis). Conversely, the Placebo group (escitalopram monotherapy) was dominated by signatures of bacterial distress, significantly enriching guanosine tetraphosphate/pentaphosphate (ppGpp) metabolism (PPGPPMET-PWY) and enterobacterial common antigen biosynthesis (PWY-7315). This microbial distress was further validated longitudinally by the functional volcano plot (**Figure 5B**), where standard SSRI monotherapy (Placebo group) preferentially accelerated pathways such as the kynurenine to NAD biosynthesis pathway. In contrast, the PS128 group demonstrated selectively up-regulated multi-acid fermentative frameworks, including acetone-isopropanol fermentation and phosphoketolase/mixed acid fermentation schemes. To deconstruct the higher-order structural organization of these predicted traits, topological co-occurrence networks were constructed (rho > 0.4, FDR < 0.05) for both cohorts. In the Probiotic group (**Figure 5C**), the interactome evolved into a highly integrated, cohesive topology organized into distinct functional modules. Within this network, *Bacteroides*, *Alistipes*, and *Parabacteroides* were mainly linked to multiple metabolism pathways, forming a prominent purple module. Concurrently, *Dietzia*, *Romboutsia*, *Agathobacter*, *Faecalibacterium*, and *Lactiplantibacillus* clustered into another coordinated unit (orange module) along with pathways governing carbohydrate metabolism, amino acid metabolism (converging onto K10670: grdA; glycine/sarcosine/betaine reductase complex component A), and transcription machinery. Meanwhile, *Bifidobacterium*, *Dorea*, *Anaerostipes*, and several other taxa were mainly linked to iron transport, mapping into a dedicated blue module. In sharp contrast, the Placebo group network (**Figure 5D**) exhibited a markedly unintegrated profile, resolving only into segregated, disjoint, and fragmented modules. This topological breakdown highlights a functional disconnection anchoring the microbial community, underscoring a potential destabilization of the gut microbiota and their associated metabolic functions under SSRI monotherapy alone. Conversely, the adjunctive PS128 treatment successfully maintained a prominently integrated, robust gut microbiota interactome, a structural reconfiguration that may critically contribute to the observed clinical symptom improvement.

**Figure 5.**
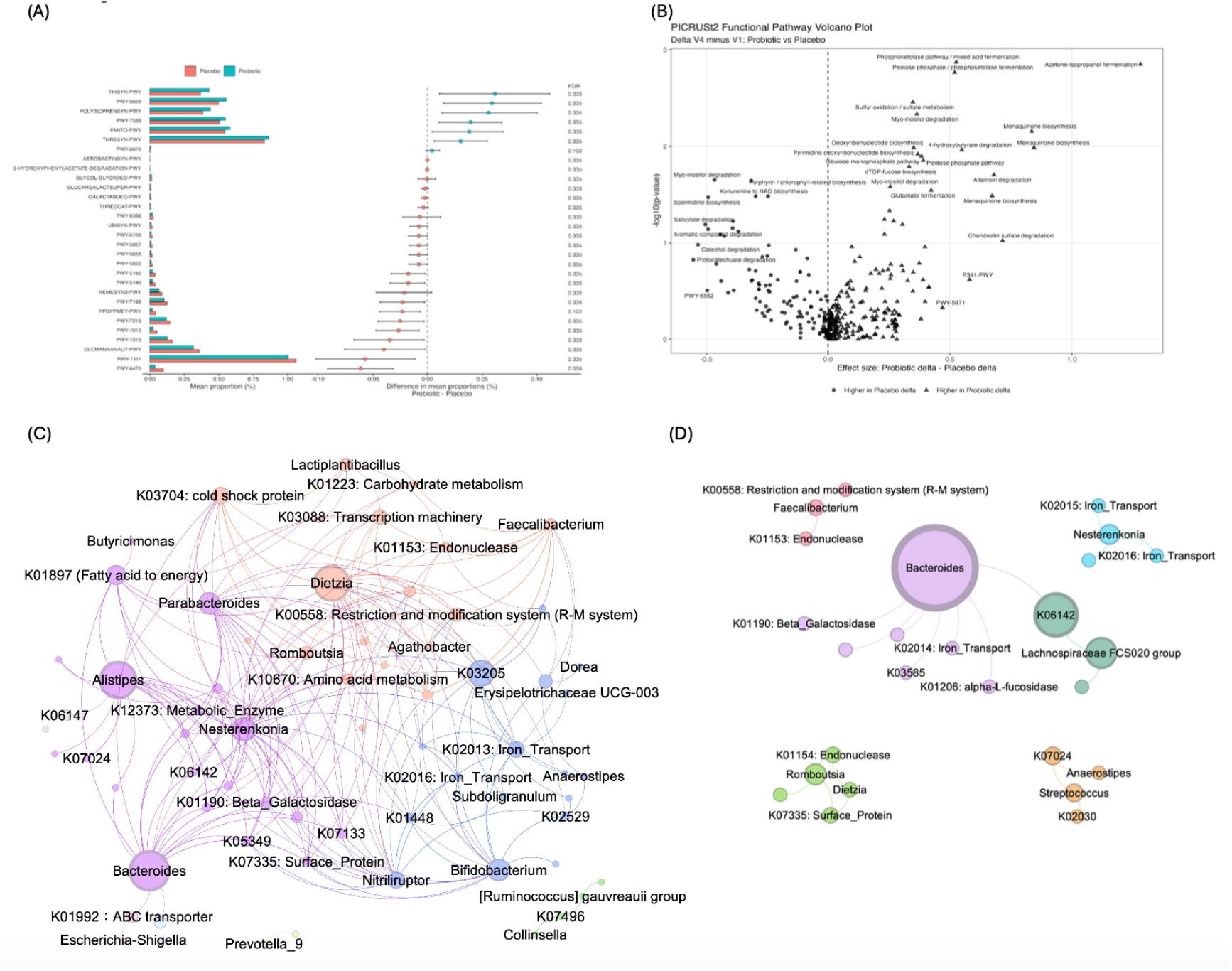
Functional metagenomic predictions and topological co-occurrence interactome networks. (A) MetaCyc pathway differentiation via PICRUSt2, with volcano plot depicting significantly upregulated and downregulated functional pathways in the PS128 group relative to placebo at Week 12. (B) Functional pathway shifts highlighting B-vitamin/cofactor biosynthesis, multi-acid fermentative frameworks, and kynurenine pathway modulation. (C, D) Higher-order topological co-occurrence networks (|rho| > 0.4, FDR < 0.05) constructed for the PS128 group (C) and placebo group (D). Nodes represent bacterial genera and predicted KEGG pathways, sized by betweenness centrality; edges denote significant Spearman correlations. Distinct functional modules were identified using the Louvain algorithm.

### 3.8. Salivary Cortisol Dynamics as a Neuroendocrine Biomarker

To investigate the neuroendocrine impact of adjunctive PS128 therapy on the hypothalamic-pituitary-adrenal (HPA) axis, salivary cortisol levels were assessed longitudinally (V1 to V4). Due to quality degradation from prolonged storage, discolored saliva samples were excluded. Because the remaining sample size was substantially smaller than that of other endpoints, these exploratory data (n = 7–12 in the PS128 group; n = 10–11 in the placebo group) are presented in the Supplementary Information (**Figure S1**). Salivary cortisol levels at baseline were comparable between groups (P > 0.05). Following the 12-week intervention, a lower salivary cortisol concentration was observed in the adjunctive PS128 group compared to the placebo group (P < 0.0001, Mann-Whitney U test; **Figure S1**). Despite the limited sample size, these preliminary findings suggest that adjunctive PS128 may contribute to modulating HPA axis reactivity in patients with MDD.

## Discussion

The clinical architecture of this randomized, double-blind, placebo-controlled trial provides crucial psychopharmacological insights into major depressive disorder (MDD) management. Since all enrolled participants were maintained on a stable first-line antidepressant regimen (escitalopram, 10 mg/day), the placebo arm mirrors the real-world trajectory of standard SSRI monotherapy, whereas the probiotic arm evaluates a dual-target, microbiota-gut-brain axis (MGBA)-mediated adjunctive strategy. Our findings demonstrate that while both cohorts exhibited standard within-group improvements—validating the established therapeutic timeline of escitalopram—the adjunctive administration of *Lactiplantibacillus plantarum* PS128 successfully broke through the traditional therapeutic ceiling of antidepressant monotherapy. This breakthrough is robustly underscored by the substantial effect sizes captured at the Week 12 endpoint (HAMD-17: Cohen’s d = –1.052; MADRS: Cohen’s d = –1.067) and a stark divergence in clinical response rates (82.8% in the PS128 group versus 25.0% in the placebo group). Furthermore, 62.1% of patients receiving adjunctive PS128 achieved full clinical remission compared to a mere 20.8% in the monotherapy arm. This profound clinical acceleration strongly implies that PS128 does not merely augment antidepressant efficacy through redundant biological pathways, but rather engages distinct, complementary microecological and metabolic mechanisms that resolve residual psychiatric symptoms.

Cross-examining these outcomes with the contemporary clinical landscape underscores the paramount importance of both intervention duration and strain specificity in psychiatric microecological therapies. Our longitudinal data demonstrated that while the clinical trajectories began to separate at Week 8, the sharpest expansion of therapeutic effect sizes materialized at Week 12. This extended temporal threshold is heavily corroborated by previous literature; short-term interventions, such as the 4-week PROVIT trial evaluating a multi-species probiotic (Reininghaus et al., 2020) and a 31-day trial utilizing an 8-strain formula (Schaub et al., 2022), frequently fail to capture stable clinician-rated symptom reductions despite successfully altering microbial composition. Conversely, clinical trials extending their timeline to 8 weeks or longer, such as the 14-strain mixture (Nikolova et al., 2023), single-strain *L. plantarum* 299v (Rudzki et al., 2019), or long-term 90-day *Bacillus coagulans* protocols (Majeed et al., 2018), consistently mirror the sustained recovery curve observed in our cohort. Biologically, this confirms that introducing psychobiotics into an environment already pharmacologically modified by chronic SSRIs requires at least 8 to 12 weeks to overcome microecological inertia. Crucially, our study deconstructs the prevailing clinical misconception that antidepressant efficacy is proportional to taxa density or cell count. While robust outcomes have been achieved using complex multi-strain consortiums (Nikolova et al., 2023), equally profound and highly targeted clinical footprints are generated by standalone single-strain configurations. For instance, single-strain *Clostridium butyricum* MIYAIRI 588 operates through specific butyrate-mediated neuroprotection (Miyaoka et al., 2018), and *Bifidobacterium longum* 1714 alleviates psychological distress (Seamans et al., 2026). By delivering a high-potency, single-strain intervention free of prebiotics or micronutrient co-factors, our findings establish that premium psychobiotic efficacy is fundamentally governed by precise, strain-specific genetic machinery rather than formulation complexity.

At the functional metagenomic level, the observed clinical breakthroughs were closely underpinned by a multi-axial taxonomic and metabolic rewiring that directly addresses the universal dysbiotic signatures of psychiatric morbidity (McGuinness et al., 2022; Nikolova et al., 2023). Our differential abundance analysis revealed a post-intervention enrichment of core short-chain fatty acid (SCFA)-producing genera in probiotic group, including *Butyricicoccus*, *Bacteroides*, Coprococcus, and *Faecalibacterium*, which historically correlate inversely with depression severity.

Intriguingly, MaAsLin2 analysis captured the significant enrichment of *Bilophila* and *Barnesiella* exclusively within the psychobiotic cohort. Rather than representing an adverse pathobiont expansion, this coordinated taxonomic shift suggests a resilient microbial reconfiguration under the selective pressure of stable escitalopram therapy (Liu et al., 2023). Within our psychobiotic cohort, this drug-induced microbial stress may uniquely neutralized by a functional bio-shield erected by the concurrent enrichment of SCFAs (especially butyrates) taxa prime the anti-inflammatory homeostatic. Furthermore, this microecological buffering is functionally mirrored by PICRUSt2 functional predictions, which demonstrated a dual-action metabolic signature. PS128 as adjunct to escitalopram drove a powerful up-regulation of neuroprotective cofactors and essential amino acids—specifically biosynthesis of vitamin B1 (THISYN-PWY), vitamin B5 (PANTO-PWY), tetrahydrofolate/vitamin B9 active form (PWY-7539), and menaquinone (Vitamin K2)—paired with a pivotal down-regulation of the neurotoxic kynurenine-to-NAD pathway (**Figures 5A** and **5B**). In severe MDD, systemic inflammation typically drives mitochondrial decay and hyperactivates the kynurenine shunt which in turn starving the central nervous system of primary monoamine building blocks (Amin et al., 2023). By generating office-obligate cofactors for mitochondrial ATP production and neurotransmitter synthesis while simultaneously muting peripheral kynurenine neurotoxic cascades via organic-acid/mixed-acid fermentations, PS128 acts as a multimodal upstream bio-factory. This creates an unassailable pharmacological-microbiological synergy: PS128 optimizes cellular energy networks and monoamine precursor availability at the source, while escitalopram maximizes synaptic serotonin persistence, thereby breaking through the therapeutic ceiling of standard monotherapy (25% response rate in the placebo group).

Beyond individual pathway alterations, macroscopic topological network analysis provided clear evidence of ecosystem-wide stabilization, offering profound physiological clarity regarding systemic neuroendocrine tuning. Under adjunctive PS128 treatment, the intestinal interactome transitioned into a highly integrated, cohesive topology organized into distinct functional modules (**Figure 5C**). The balance of neurotransmitters plays pivotal role in psychiatric disorders. Not only monoamines neurotransmitter, glycine acts as an obligatory, rate-limiting co-agonist at the binding site of central NMDA receptors, a critical venue governing synaptic plasticity and long-term potentiation that is universally blunted in treatment-resistant depression (Peyrovian et al., 2019). By centering the community network around the grdA complex, the cooperative network forged by PS128 implies a synchronized optimization of neuroactive amino acid and methyl-donor trafficking. This active cluster was dynamically anchored by a module driven by high-betweenness hubs *Bacteroides* and *Alistipes* managing ABC transporters (K01992), alongside a blue module tying *Bifidobacterium* seamlessly to specialized iron-scavenging machinery (K02013/K02016), collectively maintaining a robust, unfragmented functional interactome. In stark contrast, the placebo cohort displayed a severe architectural collapse characterized by segregated, disjoint, and fragmented sub-graphs (“isolated islands”) and profound functional decoupling (**Figure 5D**). The less diverse, fragmented microbial composition in MDD may result in dysbiosis and trigger inflammations. On the contrary, the dense, cooperative network architecture exclusively forged by PS128 adjunctive to escitalopram establishes a resilient ecological buffer that effectively quiets peripheral immune-to-brain signaling.

Although the data is not completed, the PS128 group do show a significant normalization of salivary cortisol concentrations at Week 12. By successfully down-regulating the hypothalamic secretion of corticotropin-releasing hormone and restoring the homeostatic negative-feedback set-point of the HPA axis, PS128 fundamentally shields the neuroendocrine system from the microecological side effects inherent to chronic SSRI therapy.

Despite the robust clinical outcomes and longitudinal insights generated by this trial, several methodological limitations must be acknowledged. First, while the therapeutic effect sizes within the per-protocol cohort (N = 53) were exceptionally large, the overall sample size derived from a single clinical center remains relatively modest, necessitating validation across larger, multi-center cohorts before these results can be broadly generalized to wider psychiatric populations. Second, ambulatory compliance barriers and logistical challenges resulted in a substantial rate of missing data for salivary cortisol at intermediate time points, restricting these neuroendocrine findings to a compliant subcohort presented exclusively as Supplementary Information.

Furthermore, the absence of corresponding peripheral blood samples precluded the direct measurement of systemic inflammatory cytokines or circulating neurotrophic factors. Third, regarding our microecological characterization, the taxonomy was profiled utilizing 16S rRNA gene sequencing rather than shotgun metagenomic sequencing, which inherently possesses restricted taxonomic resolution at the species level and introduces database-dependent biases during functional prediction.

Consequently, the metabolic pathways uncovered in this study—including the highly compelling modulation of multiple metabolism, monoamine precursors, and short-chain fatty acids—rely entirely on PICRUSt2 functional predictions rather than direct physical quantification. To bridge these gaps, future clinical investigations must implement deep shotgun metagenomics to achieve high-resolution strain tracking, coupled with untargeted and targeted multi-matrix (fecal, salivary, and serum) metabolomics to definitively quantify the absolute concentrations of circulating metabolites and functional profiles, thereby establishing an unassailable, quantified mechanistic bridge along the microbiota-gut-brain axis.

## Conclusion

In summary, this 12-week, randomized, double-blind, placebo-controlled trial demonstrates that adjunctive *Lactiplantibacillus plantarum* PS128 therapy successfully breaks through the therapeutic ceiling of standard SSRI monotherapy in patients with major depressive disorder. Beyond achieving superior clinician-rated symptom reductions and high clinical remission rates, adjunctive PS128 establishes a profound pharmacological-microbiological synergy with escitalopram. Mechanistically, PS128 acts as a multimodal upstream bio-factory that fortifies intestinal interactome network resilience, overexpresses vital mitochondrial and neuroprotective cofactors, and erects a taxonomic bio-shield that buffers the drug-induced microecological stress of chronic antidepressants. This systemic microecological stabilization effectively quiets peripheral immune-to-brain signaling, translating into a normalization of salivary cortisol and the homeostatic tuning of the hyperactive HPA axis. Overall, our findings position high-potency, strain-specific single psychobiotics as a transformative and clinically robust strategy to resolve residual depressive symptoms and optimize the management of psychiatric morbidity.

## Acknowledgement

The authors thank the recruited patients for their participation in this study.

## CRediT authorship contribution statement

**Yunxin Ji**: Writing – original draft, Writing – review & editing, Supervision, Resources, Project administration, Methodology, Investigation, Funding acquisition, Data curation. **Jiale Zhang**: Writing – original draft, Resources, Methodology, Investigation, Formal analysis, Data curation. **Jieqiong Hu**: Investigation. J**iaxin Mao**: Investigation. **Lanlan Wang**: Investigation. **Kuilai Wang**: Investigation. **Qingyu Zhang**: Investigation. **Zhongze Lou**: Supervision, Resources, Methodology. **Yuwei Mi**: Writing – original draft, Writing – review & editing, Supervision, Resources, Project administration, Methodology, Investigation, Funding acquisition, Data curation.

## Funding

This study was funded by Medical Science and Technology Project of Zhejiang Province (no. 2025KY1332) and Ningbo City Key R&D plan “Jie Bang Gua Shuai” (no. 2023Z197).

## Data Availability Statement

The 16S rRNA gene sequencing data generated during the current study have been deposited in the National Center for Biotechnology Information (NCBI) Sequence Read Archive (SRA) database under BioProject accession number PRJNA1509766 (https://www.ncbi.nlm.nih.gov/bioproject/PRJNA1509766). All other relevant clinical and analytical datasets supporting the conclusions of this article are available from the corresponding author upon reasonable request.

## Notes

### Competing Interest Statement

The authors have declared no competing interest.

### Clinical Trial

ChiCTR2500101411

### Author Declarations

Ethics committee/IRB of Medical Ethics Committee of the First Affiliated Hospital of Ningbo University gave ethical approval (Ningbo, China; 2023-R003-01) for this work.

